# Outcomes of percutaneous transhepatic gallbladder drainage versus percutaneous transhepatic biliary drainage for obstructive jaundice

**DOI:** 10.1101/2024.09.03.24313028

**Authors:** Tetsushi Azami, Yuichi Takano, Naoki Tamai, Jun Noda, Masataka Yamawaki, Fumitaka Niiya, Naotaka Maruoka, Fumiya Nishimoto, Akira Ishihara, Masatsugu Nagahama

## Abstract

Percutaneous transhepatic gallbladder drainage (PTGBD) is an alternative to percutaneous transhepatic biliary drainage (PTBD) for cases of obstructive jaundice in which the bile duct obstruction is below the confluence of the cystic ducts. The present study aimed to evaluate the usefulness of PTGBD and PTBD in patients with obstructive jaundice. This study enrolled patients who had undergone percutaneous biliary drainage for acute cholangitis and obstructive jaundice at two institutions between January 2017 and March 2024. Fifty-five patients were included in this analysis. However, patients with intrahepatic or hilar bile duct stenosis, post choledocholithiasis, complex cholecystitis, total bilirubin levels < 2.0 mg/dL, and uncorrectable bleeding tendency and those who had undergone the procedure and later discontinued without puncture were excluded. The technical success rates, clinical success rates, and complication rates of the procedure were evaluated. The technical success rates were 96.3% (26/27) in the PTGBD group and 82.1% (13/28) in the PTBD group. The clinical success rates were 85.2% (23/27) in the PTGBD group and 67.9% (19/28) in the PTBD group. The complication rates were 11.1% (3/27) in the PTGBD group and 17.9% (5/28) in the PTBD group. Hence, the two groups did not significantly differ in any of the endpoints. The outcomes of PTGBD were comparable to those of PTBD in patients with obstructive jaundice. Hence, PTGBD is a reasonable treatment option for cases of obstructive jaundice in which PTBD is not feasible.

## Introduction

Bile duct drainage is indicated for all patients with acute cholangitis and obstructive jaundice, except for some mild cases that improve with antimicrobial therapy alone. The Tokyo Guidelines 2018 recommend endoscopic biliary drainage (EBD) as the first-line treatment for bile duct drainage [1]. However, in some cases, EBD cannot be performed due to factors such as the patient’s general condition, postoperative reconstructed intestinal tract, and gastrointestinal stenosis. In recent years, endoscopic ultrasound-guided biliary drainage (EUS-BD) has been the salvage procedure of choice for such cases. If EBD and EUS-BD are challenging to perform, percutaneous transhepatic biliary drainage (PTBD) is considered. However, its technical success rate is 63% in patients with nondilated bile ducts and 86% in patients with dilated ducts, which is not sufficient [2].

Alternatively, percutaneous transhepatic gallbladder drainage (PTGBD) is a drainage technique for acute cholecystitis. It has a high success rate and can be useful for bile duct drainage if the bile duct obstruction is downstream of the cystic duct. EUS-guided gallbladder drainage has been reported to be useful for EUS-guided hepaticogastrostomy failure in obstructive jaundice [3]. PTGBD can serve as an alternative treatment strategy to PTBD. A case series and a small number of retrospective cohort studies have evaluated the use of PTGBD for cholangitis and obstructive jaundice [4-7]. However, to the best of our knowledge, no studies have compared PTGBD and PTBD. The present study aimed to examine the usefulness of PTGBD and PTBD in patients with obstructive jaundice.

## Materials and Methods

### Study design and participants

This retrospective study was based on an analysis of data collected from the patients’ electronic medical records. Patients who had undergone percutaneous biliary drainage for acute cholangitis and obstructive jaundice at Showa University Fujigaoka Hospital or Hitachi Medical Center between January 1, 2017 and March 31, 2024 were included in this analysis. Patients with intrahepatic or hilar bile duct stenosis, hepaticojejunostomy anastomotic stricture, coexisting cholecystitis, total bilirubin (T.bil) level < 2.0 mg/dL, and uncorrectable bleeding tendency and those who had undergone the procedure and later discontinued without puncture were excluded.

The original data were accessed for research purposes on August 16, 2024. The study was conducted according to the Declaration of Helsinki guidelines and was approved by the Showa University Research Ethics Review Board (Approval Number: 2024-106-B). Due to the retrospective nature of the study, the need for informed consent was waived by the IRB.

### Outcomes

The primary endpoints were technical success rates, clinical success rates, and complication rates. Technical success was defined as catheter placement into the biliary tract. Clinical success was defined as a decrease in T.bil levels to <2.0 mg/dL or a 50% decrease in T.bil levels after 2 weeks. Complications were evaluated based on the Clavien–Dindo classification [8]. The severity of cholangitis was also assessed using the Tokyo Guidelines 2018 [1].

### Procedure details

For percutaneous drainage, PTBD was the first choice, but in cases with poor intrahepatic bile duct dilatation or when bile duct puncture was difficult due to respiratory fluctuations, PTGBD was selected at the surgeon’s discretion. PTGBD was performed using a one-step technique (Fig 1). PTBD was performed with either one- or two-step techniques based on the surgeon’s discretion (Fig 1). If necessary, analgesics (pentazocine 15 mg) were administered intravenously before puncture.

**Fig 1.**
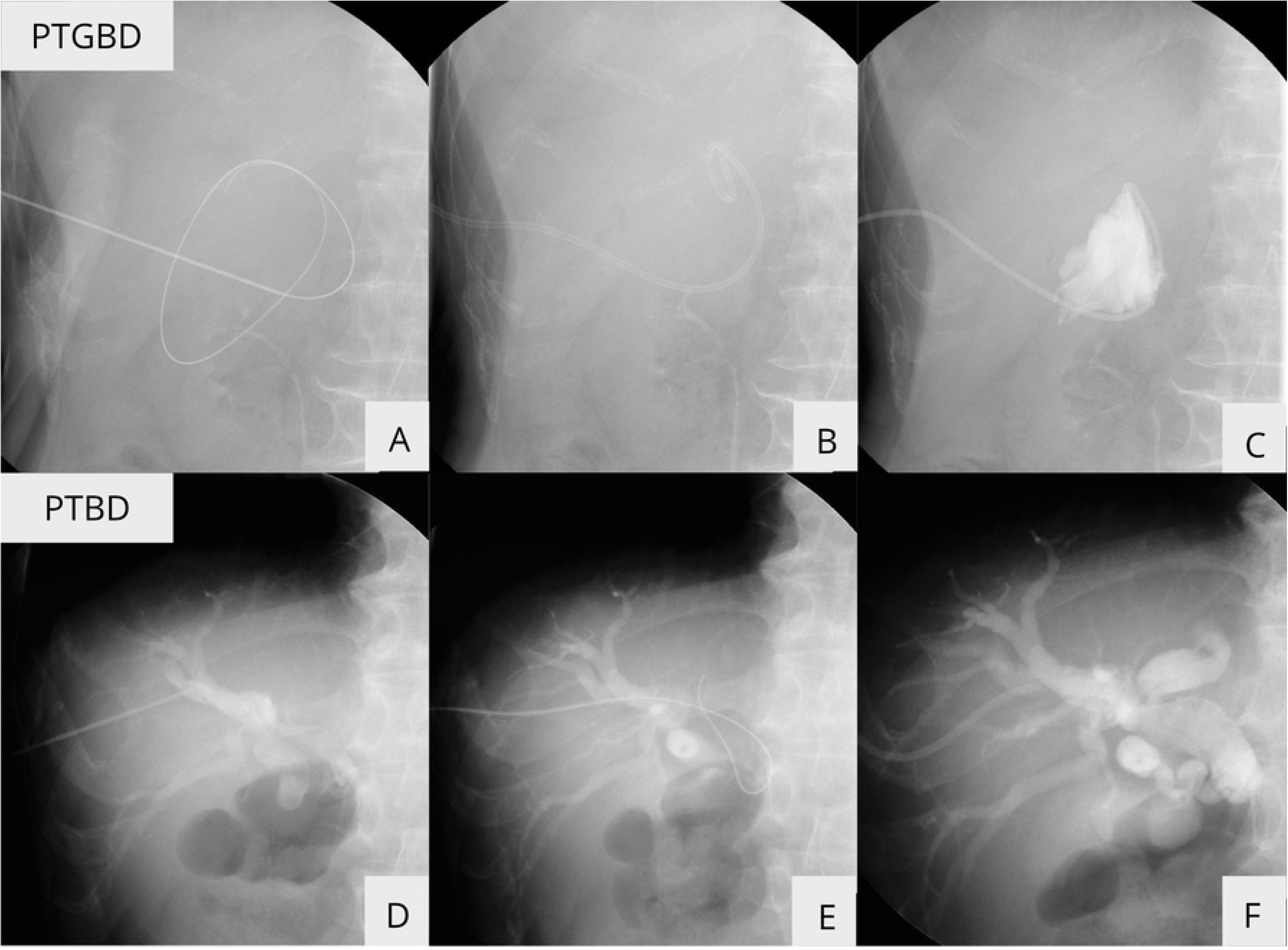
Percutaneous transhepatic gallbladder drainage (PTGBD) and percutaneous transhepatic biliary drainage (PTBD) procedure. (A) 18-G needle was inserted into the gallbladder. Next, a 0.035-in guide wire was placed in the gallbladder. (B) 8-Fr pigtail catheter was inserted. (C) Cholangiography was performed. (D) 18-G needle was inserted into the gallbladder and cholangiography was performed. (E) A 0.035-in guide wire was placed in the bile duct. (F) 8-Fr pigtail catheter was inserted.

### One-step method

After administering local anesthesia with lidocaine, an 18-G needle (Hanaco Medical, Saitama, Japan) was inserted into the gallbladder or bile duct under ultrasound guidance, and a small amount of bile was aspirated. Next, a 0.035-in guide wire (Radifocus, Terumo, Tokyo, Japan) was placed in the gallbladder or bile duct under fluoroscopy. After a small skin incision was made and dilated with a dilator, a 6–8-Fr pigtail catheter (UreSil, L.C.C., Illinois, the USA) was inserted.

### Two-step method

After administering local anesthesia with lidocaine, a 20-G needle was inserted under ultrasound guidance. A small amount of bile was aspirated. Then, cholangiography was conducted (Urografine 60%, Byer AG, Nordrhein-Westfalen, Germany). Next, a 0.018-in guide wire was placed in the bile duct under fluoroscopic guidance. A small skin incision was made, an introducer set was inserted, and all but the sheath was removed. Then, a 0.035-in guide wire was inserted, the sheath was removed, and the bile duct was dilated with a dilator. Finally, a 7-Fr pigtail catheter (CLINY PTCD Kit, Create Medic Co., Yokohama, Japan) was implanted.

### Statistical analysis

The outcomes of the PTGBD and PTBD groups were compared using the Mann– Whitney U test for continuous variables and chi-square test or Fisher’s exact test for nominal variables. A p value of <0.05 was considered statistically significant. R version 4.0.3 (R Foundation for Statistical Computing, Vienna, Austria) was used for data analysis.

## Results

### Clinical characteristics of the patients

Percutaneous drainage was performed on 79 patients with cholangitis and obstructive jaundice between January 1, 2017, and March 31, 2024. Among them, two presented with hilar bile duct stenosis, seven with hepaticojejunostomy anastomotic stricture, seven with coexisting cholecystitis, and eight with T.bil levels < 2.0 mg/dL were excluded. Hence, 55 patients were finally included in this study.

Of 55 patients, 27 and 28 underwent PTGBD and PTBD, respectively. One patient in the PTGBD group and five in the PTBD group experienced technical failure. Technical success was 26 in the PTGBD group and 23 in the PTBD group (21 one-step and 2 two-step procedures) (Fig 2).

**Fig 2.**
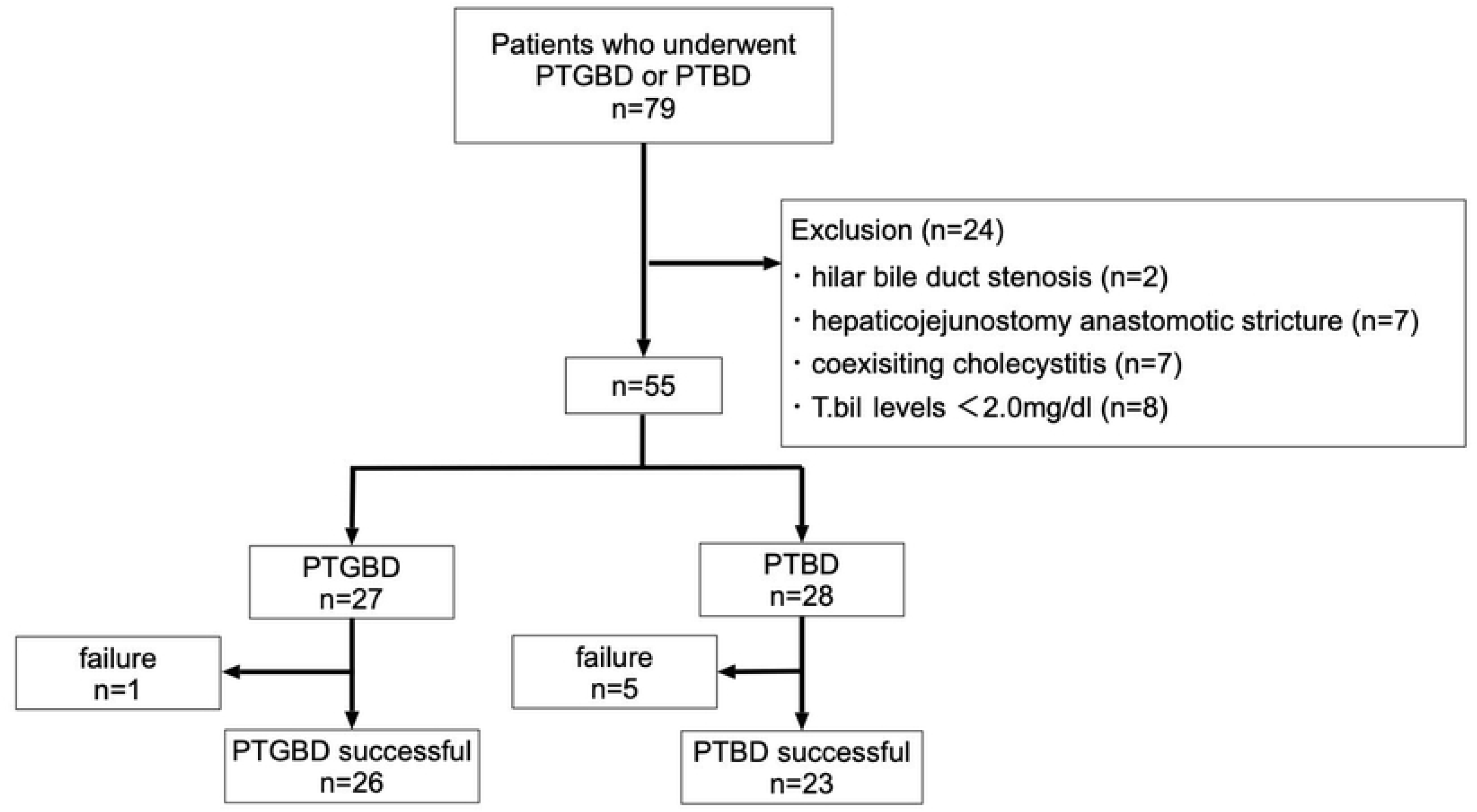
Participant flow chart. PTGBD: percutaneous transhepatic gallbladder drainage. PTBD: percutaneous transhepatic biliary drainage.

In terms of technical failures, in one patient, PTGBD was unsuccessful because the dilator could not be inserted due to the lack of gallbladder distension. Surgical treatment was then performed. In five patients in whom PTBD failed, three and two underwent PTGBD and EUS-BD, respectively (Fig 2). Table 1 shows the characteristics of the patients. The PTGBD and PTBD groups did not significantly differ in terms of age, sex, and use of antithrombotic medication. The median Charlson Comorbidity Index (CCI) values were 7 (3–12) in the PTGBD group and 9 (4–11) in the PTBD group. The PTBD group (9.33 [2.0–26.2] mg/dL) had significantly higher baseline T.bil levels than the PTGBD group (4.30 [2.3–22.7] mg/dL). Severe cholangitis was significantly more common in the PTGBD group than in the PTBD group (14 [51.9%] vs 5 [17.9%]). In terms of obstruction etiology, 18 (66.7%) and 9 (33.3%) patients in the PTGBD group presented with benign and malignant diseases, respectively.

**Table 1.**
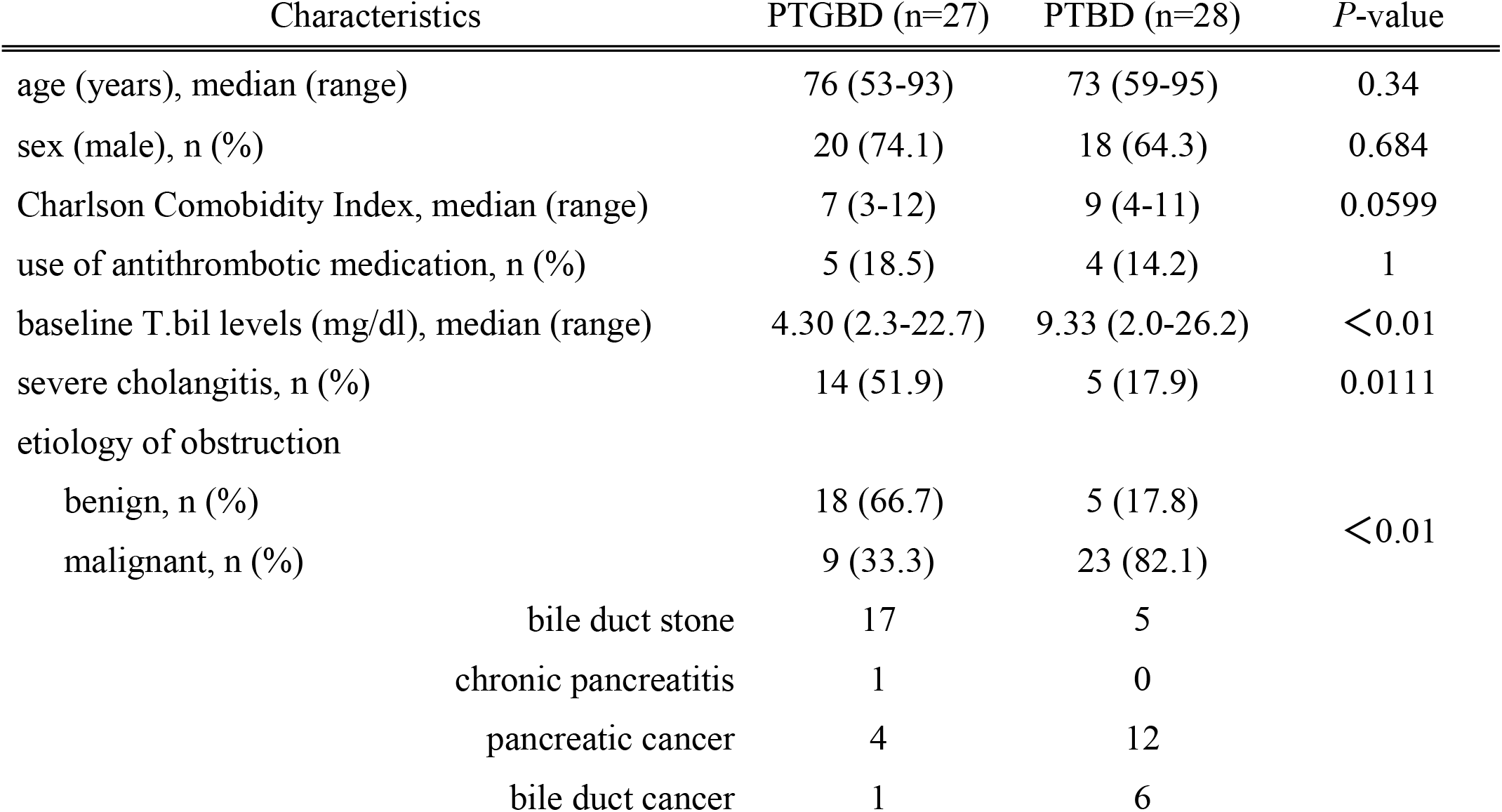
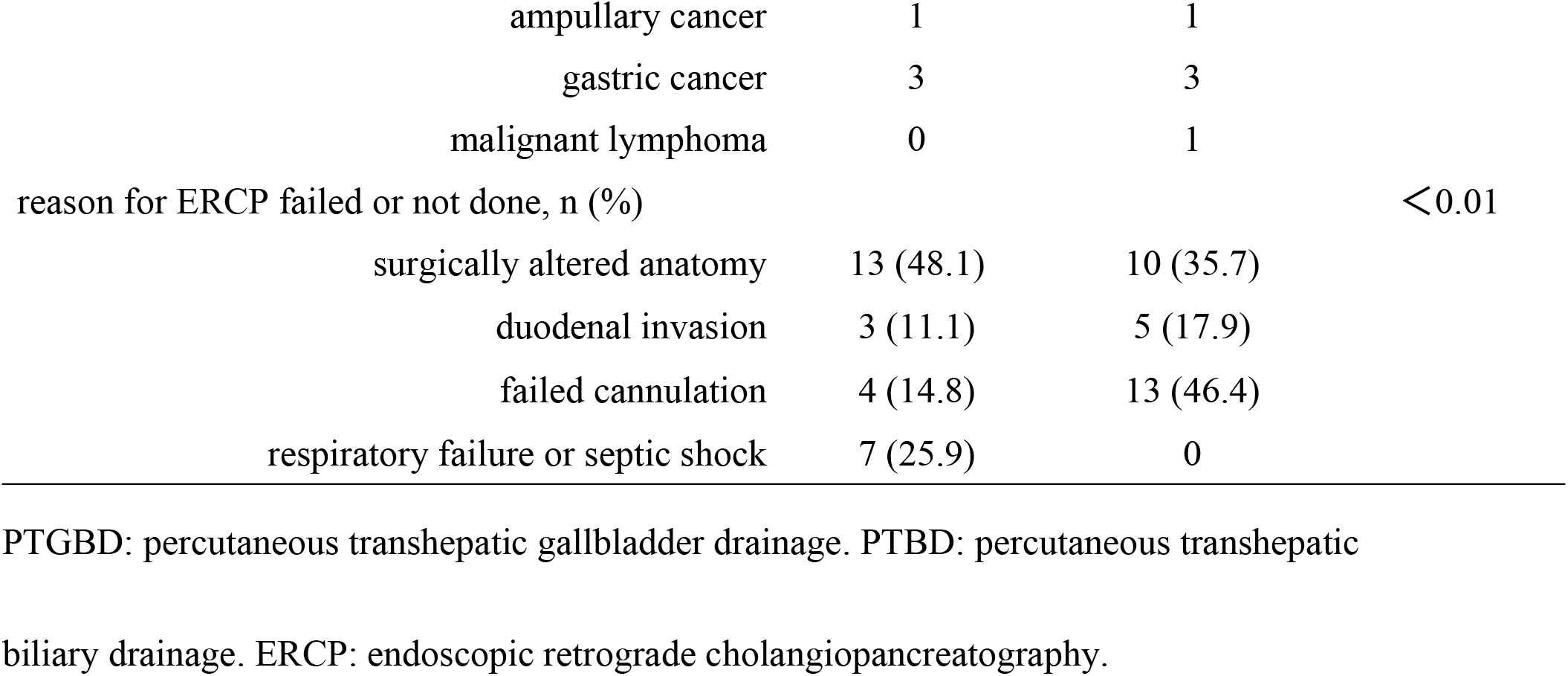
Comparison of characteristics between percutaneous transhepatic gallbladder drainage and percutaneous transhepatic biliary drainage.

Moreover, 5 (17.8%) and 23 (82.1%) patients in the PTBD group presented with benign and malignant diseases, respectively. Hence, benign disease was more common in the PTGBD group than in the PTBD group. The most common reasons for failure or absence of endoscopic retrograde cholangiopancreatography (ERCP) were postoperative surgically altered anatomy in the PTGBD group (n = 13 [48.1%]) and failed cannulation in the PTBD group (n = 13 [46.4%]) (Table 1).

### Outcomes

PTGBD: percutaneous transhepatic gallbladder drainage. PTBD: percutaneous transhepatic biliary drainage.

The technical success rates were 96.3% (26/27) in the PTGBD group and 82.1% (23/28) in the PTBD group. The clinical success rates were 85.2% (23/27) in the PTGBD group and 67.9% (19/28) in the PTBD group. Hence, the two groups did not significantly differ in terms of technical success and clinical success rates. Although not included in the PTBD group, one patient who did not achieve a clinical success to PTGBD achieved a clinical success to PTBD. The complication rates were 11.1% (3/27) in the PTGBD group and 17.9% (5/28) in the PTBD group. Thus, there were no significant differences in terms of the complication rates between the two groups (Table 2).

**Table 2.**
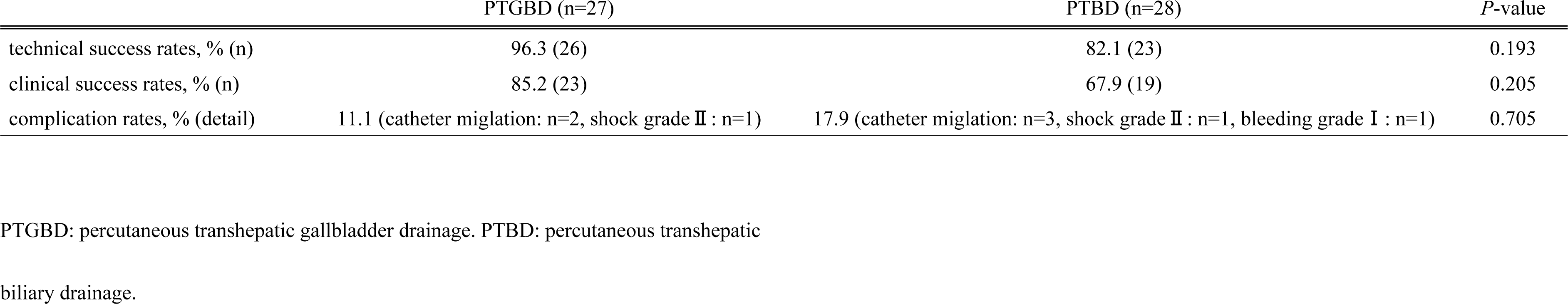
Comparison of outcomes between Percutaneous transhepatic gallbladder drainage and percutaneous transhepatic biliary drainage.

## Discussion

Cholecystocentesis, which was first reported by Burckhardt and Muller in 1921, has a long history [9]. Subsequently, Huard and Du-Xuan-Hop et al. successfully performed intrahepatic bile duct puncture in 1937 [10]. The introduction of echo-guided puncture and improvements in treatment tools led to therapeutic application of percutaneous biliary puncture. PTBD was first reported by Glenn et al. in 1962 [11]. PTGBD was first reported by Elyaderani and Gabriele in 1979 [12] and performed by Radder in 1982 in patients with acute cholecystitis [13]. To date, in the Tokyo Guidelines, PTGBD is recommended as the standard drainage method for patients with cholecystitis who are at high risk for surgery, and PTBD is an alternative treatment to EBD in patients with cholangitis [1].

In recent years, EUS-BD has been gaining popularity as a biliary drainage technique since the study of Giovannini [14]. Moreover, EUS-GBD has been reported to be useful in treating obstructive jaundice in cases of EUS-guided hepaticogastrostomy failure if the bile duct obstruction was downstream of the confluence of the cystic duct. Furthermore, PTGBD may be useful in obstructive jaundice. A case series and a few retrospective cohort studies have long documented the efficacy of PTGBD for obstructive jaundice [4-7].

Li et al. and Park et al. reported that PTGBD resulted in jaundice control in 91% (29 / 32) and 100% (20 / 20) of patients with obstructive jaundice. Only 3% (one patient with biliary peritonitis) and 5% (one patient with catheter migration) of patients presented with complications, respectively. And technical success was 100% in both studies. Both studies have revealed that PTGBD is useful in treating obstructive jaundice [4, 5].

No reports have compared PTGBD and PTBD. This is the first report comparing the efficacy of PTGBD and PTBD in obstructive jaundice. The technical success, clinical success, and complication rates of PTGBD and PTBD were comparable in this study.

PTGBD for obstructive jaundice should be considered as an alternative treatment to PTBD. However, in one case, PTBD improved jaundice in a patient who showed no clinical response to PTGBD. Treatment via the percutaneous drainage route (rendezvous procedure, percutaneous bile duct stenting, and stone removal) is also less difficult with PTBD than with PTGBD. In patients considering treatment via the percutaneous drainage route, PTBD can be a better option because of the ease of subsequent procedures.

Although there were no significant differences in terms of outcomes between the two groups, the clinical characteristics of the two groups differed. In terms of disease etiology, benign diseases such as common bile duct stones were more common in the PTGBD group than in the PTBD group. Meanwhile, malignant diseases were more common in the PTBD group than in the PTGBD group. Severe cholangitis was significantly more common in the PTGBD group than in the PTBD group. Severe cholangitis caused by bile duct stones requires a short treatment time due to unstable vital signs. In several cases, it is difficult to obtain patient cooperation and respiratory arrest at the time of puncture. Therefore, PTGBD, which is less difficult to perform, was selected. By contrast, obstructive jaundice caused by malignant diseases is generally more severe than that caused by benign disease. However, it is less likely to be complicated by severe cholangitis. The high degree of bile duct dilatation and the ease of patient cooperation might have been the reasons for selecting PTBD. The PTBD group can have a higher CCI than the PTGBD group because of the higher number of patients with malignant diseases. There were no statistically significant differences. However, it should not be underestimated that the PTGBD group was more likely to have a higher technical success rate and a lower incidence rate of accidents.

The greatest advantage of PTGBD is its ease of use and high technical success rate. PTGBD and PTBD should be used interchangeably in different cases. However, in cases where PTBD is challenging to perform, PTGBD should be considered as an alternative treatment.

The present study had several limitations. First, it was a retrospective study conducted at two centers, and the number of cases was small. The two groups differed in terms of clinical characteristics, with the PTGBD group having milder jaundice. The type of procedure might have been influenced by the operator’s subjective bias. Future studies comparing PTBD and PTGBD under identical conditions should be performed.

## Conclusions

The outcomes of PTGBD are comparable with those of PTBD in patients with obstructive jaundice. Thus, PTGBD is a reasonable treatment option in cases of obstructive jaundice in which the intrahepatic bile duct dilatation is poor or PTBD is challenging to perform due to the patient’s general condition.

## Data Availability

All relevant data are within the manuscript and its Supporting Information files.

## Acknowledgments

None.

## References

[1] Yokoe M, Hata J, Takada T, Strasberg SM, Asbun HJ, Wakabayashi G, et al. Tokyo Guidelines 2018: diagnostic criteria and severity grading of acute cholecystitis (with videos). J Hepatobiliary Pancreat Sci. 2018;25(1):41–54.

[2] Saad WE, Wallace MJ, Wojak JC, Kundu S, Cardella JF. Quality improvement guidelines for percutaneous transhepatic cholangiography, biliary drainage, and percutaneous cholecystostomy. J Vasc Interv Radiol. 2010;21(6):789–95.

[3] Imai H, Kitano M, Omoto S, Kadosaka K, Kamata K, Miyata T, et al. EUS-guided gallbladder drainage for rescue treatment of malignant distal biliary obstruction after unsuccessful ERCP. Gastrointest Endosc. 2016;84:147–151.

[4] Park JM, Kang CD, Lee M, Park SC, Lee SJ, Jeon YH, et al. Percutaneous cholecystostomy for biliary decompression in patients with cholangitis and pancreatitis. J Int Med Res. 2018;46(10):4120–4128.

[5] Li YL, Wong KH, Chiu KW, Cheng AK, Cheung RK, Yam MK, et al. Percutaneous cholecystostomy for high-risk patients with acute cholangitis. Medicine (Baltimore). 2018;97(19):e0735.

[6] Ren Z, Xu Y, Zhu S. Percutaneous transhepatic cholecystostomy for choledocholithiasis with acute cholangitis in high-risk patients. Hepatogastroenterology. 2012;59:329–331.

[7] Shitrit AB-G, Braverman D. Interval percutaneous cholecystostomy is effective for decompression of the common bile duct in high-risk elderly patients prior to endoscopic retrograde cholangiopancreatography. Gerontology. 2008;54:144–147.

[8] Dindo D, Demartines N, Clavien PA. Classification of surgical complications: a new proposal with evaluation in a cohort of 6336 patients and results of a survey. Ann Surg. 2004;240(2):205–13.

[9] Burckhardt H, Muller W. Study on the puncture of the gallbladder and their X-ray imaging. Dtsch Z Chir. 1921;162:168–197.

[10] Huard P, Du-Xuan-Hop. Transhepatic bile duct puncture. Bull Soc Med Chir Indochine. 1937;15:1090–1110.

[11] Glenn F, Evans JA, Mujahed Z, Thorbjarrnarson B. Percutaneous transhepatic cholangiography. Ann Surg. 1962;156:451–460.

[12] Elyaderani M, Gabriele OF. Percutaneous cholecystostomy and cholangiography in patients with obstructive jaundice. Radiology. 1979;130:601–602.

[13] Radder RW. Percutaneous cholecystostomy. AJR Am J Roentgenol. 1982;139:1240–1241.

[14] Giovannini M, Moutardier V, Pesenti C, Bories E, Lelong B, Delpero JR. Endoscopic ultrasound-guided bilioduodenal anastomosis: a new technique for biliary drainage. Endoscopy. 2001;33:898–900.

